# Pre-symptomatic transmission of SARS-CoV-2 infection: a secondary analysis using published data

**DOI:** 10.1101/2020.05.08.20094870

**Authors:** Miriam Casey, John Griffin, Conor G. McAloon, Andrew W. Byrne, Jamie M Madden, David Mc Evoy, Áine B. Collins, Kevin Hunt, Ann Barber, Francis Butler, Elizabeth A. Lane, Kirsty O’Brien, Patrick Wall, Kieran A. Walsh, Simon J. More

## Abstract

**Objective:** To estimate the proportion of pre-symptomatic transmission of SARS-CoV-2 infection that can occur and timing of transmission relative to symptom onset.

**Setting/design:** Secondary analysis of international published data.

**Data sources:** Meta-analysis of COVID-19 incubation period and a rapid systematic review of serial interval and generation time, which are published separately.

**Participants:** Studies were selected for analysis if they had transparent methods and data sources and they provided enough information to simulate full distributions of serial interval or generation time. Twenty-three estimates of serial interval and five of generation time from 17 publications were included.

**Methods:** Simulations were generated of incubation period and of serial interval or generation time. From these, transmission times relative to symptom onset were calculated and the proportion of pre-symptomatic transmission was estimated.

**Outcome measures:** Transmission time of SARS-CoV-2 relative to symptom onset and proportion of pre-symptomatic transmission.

**Results:** Transmission time ranged from a mean of 2.91 (95% CI: 3.18-2.64) days before symptom onset to 1.20 (0.86-1.55) days after symptom onset. Unweighted pooling of estimates of transmission time based on serial interval resulted in a mean of 0.60 days before symptom onset (3.01 days before to 1.81 days after). Proportion of pre-symptomatic transmission ranged from 42.8% (39.8%-45.9%) to 80.6% (78.1%-83.0%). The proportion of pre-symptomatic transmission from pooled estimates was 56.4% (34.9%-78.0%).

**Conclusions:** Whilst contact rates between symptomatic infectious and susceptible people are likely to influence the proportion of pre-symptomatic transmission, there is substantial potential for pre-symptomatic transmission of SARS-CoV-2 in a range of different contexts. Our work suggests that transmission is most likely in the day before symptom onset whereas estimates suggesting most pre-symptomatic transmission highlighted mean transmission times almost three days before symptom onset. This highlights the need for rapid case detection, contact tracing and quarantine.

**Strengths and weaknesses of this study:**

- We estimate the extent and variation of pre-symptomatic transmission of SARS-CoV-2 infection across a range of contexts. This provides important information for development and targeting of control policies and for the parameterisation of transmission models.
- This is a secondary analysis using simulations based on published data, some of which is in pre-print form and not yet peer-reviewed. There is overlap in the contact tracing data that informed some of our source publications. We partially address this by summarising data at source location level as well as at study level.
- Populations where symptomatic people are rapidly isolated are likely have relatively more pre-symptomatic transmission. This should be borne in mind whilst interpreting our results, but does not affect our finding that there is substantial potential for pre-symptomatic transmission of SARS-CoV-2 infection.
- A strength of our approach is that it builds an understanding of pre-symptomatic transmission from a range of estimates in the literature, facilitates discussion for the drivers of variation between them, and highlights the consistent message that consideration of pre-symptomatic transmission is critical for COVID-19 control policy.

## Introduction

There is currently a pandemic of coronavirus disease (COVID-19), a recently emerged and rapidly spreading infectious disease that is caused by the novel coronavirus, severe acute respiratory syndrome coronavirus 2 (SARS-CoV-2). There are large direct impacts of COVID-19 amongst known cases. As of 2^nd^ of June 2020, the World Health Organization has reported 6, 194, 533 confirmed cases and 376, 320 deaths due to COVID-19 [1]. In China, 14% and 5% of cases were classified as severe and critical, respectively [2], and a report from Italy showed that 18% of cases required intensive care [3]. There are also major indirect impacts of COVID-19 and its control measures on other aspects of health care [4-6] and on the economy [7,8].

As there is currently no COVID-19 vaccine ready for widespread use, primary control measures entail reducing transmission from infectious individuals. These include case isolation, contact tracing and quarantine, physical distancing and hygiene measures [9]. Infectious people are predominantly identified by reported symptoms of COVID-19.

In absence of active surveillance, infectious people without symptoms may not be quarantined, and therefore may have more contacts with susceptible people resulting in increased COVID-19 transmission. Therefore, quantifying the transmission potential of COVID-19 before or in the absence of symptoms will inform disease control measures and predictions of epidemic progression.

Characteristics of pre-symptomatic and asymptomatic transmission are potentially different, and separate approaches may be required to understand them. We aimed to capitalise upon the considerable information about pre-symptomatic transmission that can be inferred from contact tracing studies. Therefore, we focus here on transmission from people before they develop symptoms rather than that from people who never develop symptoms. This addresses the urgent need for more data on extent of pre-symptomatic transmission which has been highlighted by those developing models to inform policies [10].

The pre-symptomatic transmission potential of COVID-19 has been highlighted by case reports [11-20]. The potential for pre-symptomatic transmission was also suggested by detection of viral genome in upper respiratory samples prior to symptoms [21-23].

Live virus has been isolated very soon after symptom onset [24]. These findings are supported by quantitative studies based on contact tracing, reporting serial intervals or generation times similar in duration or shorter than incubation periods in some situations [25-28], and even evidence of symptoms manifesting in the infectee prior to the infector in some cases [28-31]. Several studies have quantified the proportion [25,27-29,32] and timing [25,28,29,32] of pre-symptomatic transmission, using a variety of datasets and methodologies.

Here, using secondary analysis of data collated in meta-analysis [33] and a rapid systematic review [34] that are published separately, we apply a standardised methodology to estimate the proportion and timing of pre-symptomatic transmission of COVID-19 in a range of different contexts.

## Methods

### Principles of methodology

If generation time, the duration in days between time of infection of a secondary case (infectee) and that of its primary case (infector), is longer than incubation period, the time between infection and symptom onset in the infector, transmission will have occurred after symptom onset (Scenario A in Figure 1). If generation time is shorter than incubation period, pre-symptomatic transmission will have occurred (Scenarios B and C in Figure 1). If an infector and infectee incubation periods are taken to be independent and identically distributed, serial interval, the time between infector and infectee symptom onset, can be taken as an approximation of generation time [35,36], although serial interval will have more variation [26]. Table 1 contains definitions relevant to our analysis.

**Figure 1:**
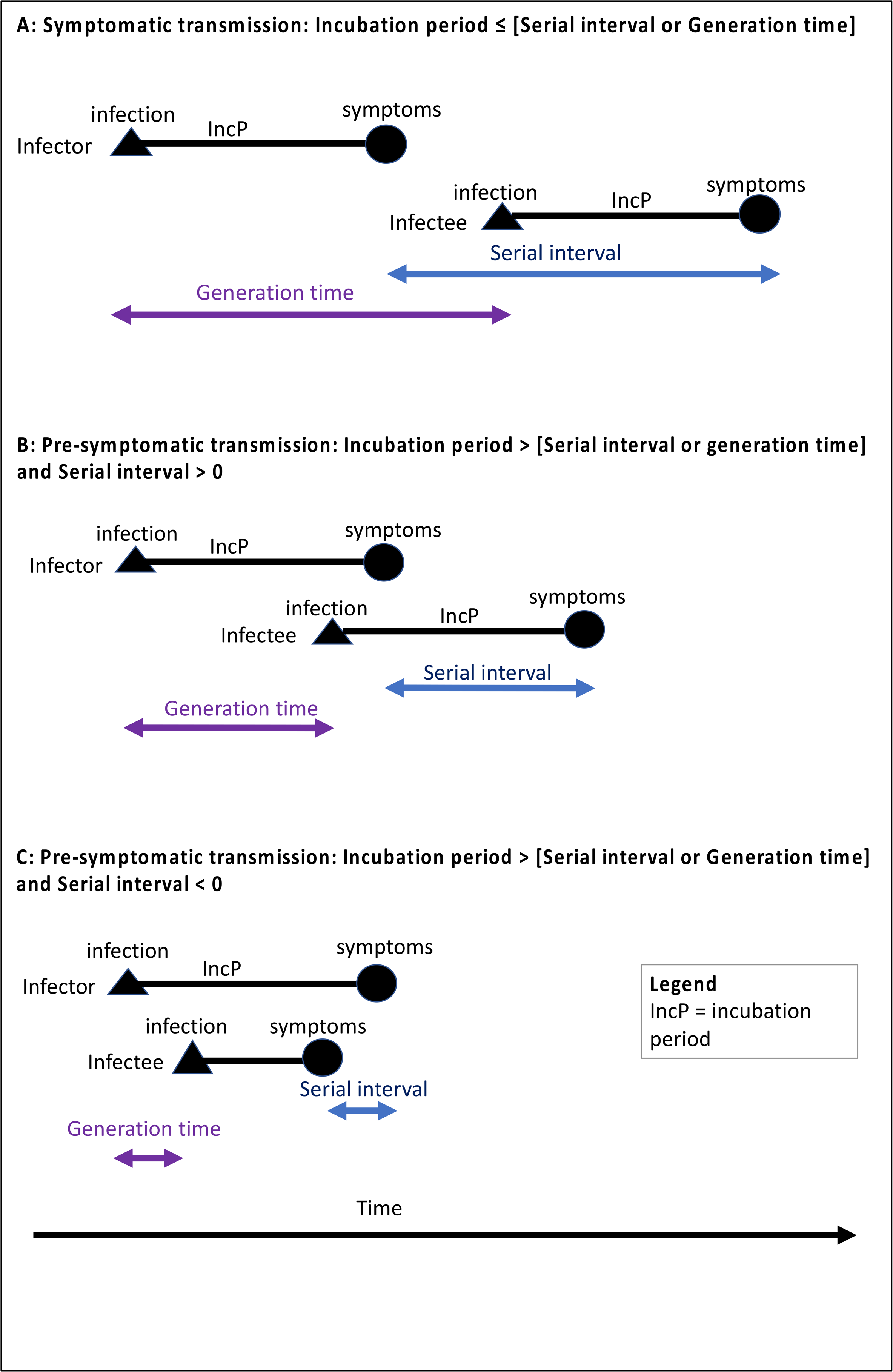
Schematic illustration of incubation period, generation time and serial interval at infector-infectee pair level. Scenario A: if serial interval/generation time is longer than incubation period, transmission occurs after symptom onset. Scenario B: if serial interval/generation time is shorter than incubation period, transmission occurs prior to symptom onset. Scenario C: A negative serial interval is possible if symptoms manifest in the infectee before the infector. If incubation period is assumed to be independent and identically distributed, mean serial interval will approximate mean generation time.

**Table 1:** Definitions referred to in this review.

|  |  |
| --- | --- |
| **Asymptomatic** | An infected person who never develops symptoms of the disease |
| **Pre-symptomatic** | An infected person before they develop symptoms of the disease. |
| **Duration of infectiousness** | The time interval in days during which an infectious agent may be transferred directly or indirectly from an infected person to another person [59]. |
| **Incubation period** | The time interval in days between invasion by an infectious agent and appearance of the first signs or symptoms of the disease in question [59]. |
| **Serial interval** | The duration in days between symptom onset of a secondary case and that of its primary case. |
| **Generation time or generation interval** | The duration in days between time of infection of a secondary case (infectee) and that of its primary case (infecter). |
| **Transmission pair** | An infected person (infecter) and a person who they transmit the pathogen to (infectee). |
| **Latent period** | The period from the point of infection to the beginning of the state of infectiousness [60]. This period corresponds to the “exposed” (E) compartment of a susceptible-exposed-infectious-recovered/removed (SEIR) model. |
| **Transmission time relative to symptom onset** | The time of transmission of an infectious agent from an infecter to an infectee in days relative to the onset of symptoms in the infecter. |
| **Proportion of pre-symptomatic transmission** | The proportion of all transmission events that occur before the onset of symptoms in the infecter. |

### Meta-analysis and rapid systematic review

Data about incubation period, serial interval and generation time were sourced through on our separately published metanalysis of incubation period [33] and rapid systematic review of serial interval and generation time [34]. As described fully elsewhere [33,34], literature searches covered publication dates between the 1^st^ of December 2019 and 8^th^ of April 2020 for incubation period, extending to the 27^th^ of April 2020 for generation time / serial interval. A dedicated team searched for publications on the electronic databases PubMed [37], Google Scholar [38], MedRxiv [39] and BioRxiv [40] with the following keywords: “Novel coronavirus” OR “SARS-CoV-2” OR “2019-nCoV” OR “COVID-19” AND “serial interval” OR “latent period” OR “incubation period” OR “generation time” OR “infectiousness” OR “pre-symptomatic” OR “asymptomatic”). The dynamic curated PubMed database “LitCovid” [41,42] was also monitored. In addition, publicly available reports from the World Health Organization [1,9,43], European Centre for Disease Prevention and Control [44] and Centres for Disease Control and Prevention Morbidity and Mortality Weekly Reports [45] were monitored, as well as curated summaries on relevant topics from the American Association for the Advancement of Science [46] and the Nature Journal [47]. Both our meta-analysis [33] and rapid systematic review [34] completed checklists to show fulfilment of Preferred Reporting Items for Systematic reviews and Meta-Analyses - extension for Scoping Reviews (PRISMA-ScR) [48].

Based on the estimates reported by our meta-analysis [33] and rapid systematic review [34], we simulated data for incubation period, serial interval and generation time. We subtracted incubation period from serial interval or generation time to infer transmission time relative to onset of symptoms and to estimate the proportion of pre-symptomatic transmission.

### Data extraction

All analyses were conducted in the R statistical environment [49]. For each publication included in our analyses, parameters describing the distributions of generation time or serial interval, and the location and dates of collection of the contact tracing data from which they were generated were collated.

If not reported directly, gamma shape and rate parameters were calculated from mean and standard deviation, either directly or using the “epitrix” [50] package as follows.

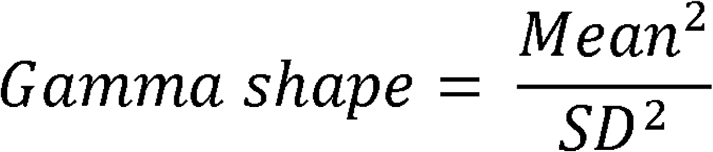

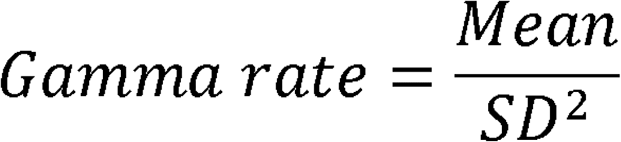

If lognormal distribution parameters (Meanlog and SDlog) were not directly reported, they were estimated from the reported mean and standard deviation (SD) as follows:

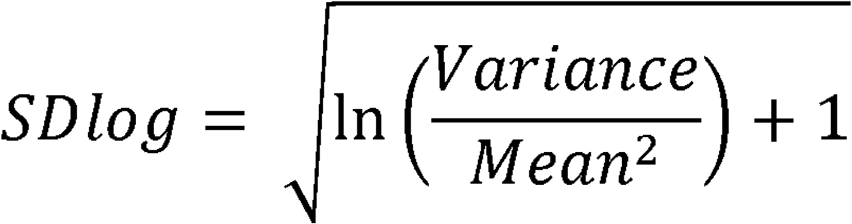

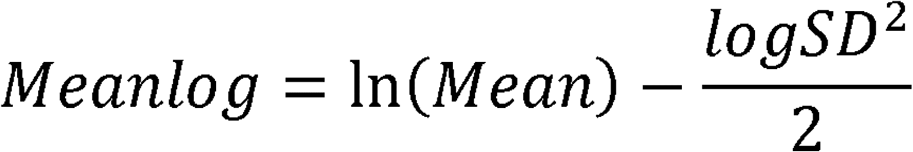

Weibull parameters were estimated from the reported mean and standard deviation using the “ “mixdist” [51] R package.

The incubation period that we used from the meta-analysis [33] had a lognormal distribution (Mean = 5.8 (95% CI (5.01, 6.69 days). It, Median = 5.1 (4.5, 5.8) days,, SD = 3 (,2.68-3.34), Meanlog = 1.63, (1.51, 1.75) and SDlog = 0.5 (0.45, 0.55) respectively.).

As serial interval has more variation than generation time [26], we considered them separately for plotting and summary purposes. For the two studies [26,27] that estimated generation time, we also generated serial interval simulations to allow a more direct comparison with the other estimates based on serial interval. One of these, [26], related serial interval to generation time with the following approach: Serial interval of an infectee can be expressed as generation time of the infectee plus the difference between the incubation periods of the infectee and the infector (Figure 1). That is, the incubation period used for the generation time estimation was simulated twice to generate two samples (“inc 1” and “inc2”). The extra variation in serial interval compared to generation time was then simulated by:

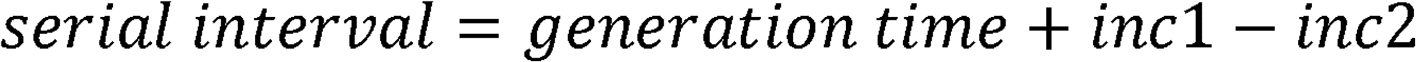

We repeated this estimation to simulate the serial interval for study [26] and cross-checked the simulation against the summary statistics were reported in that publication. We then estimated a serial interval from the generation time of study [27], using the same methodology and the same incubation period as was used to infer generation time in study [27].

One further study [52] did not directly supply enough information to simulate a distribution, but supplied their code and data from which they estimated the serial interval. For this study, we fitted distributions to their data by maximum likelihood estimation with the “fitdistrplus” package [53] and chose one based on AIC values and visual cross-checking by plotting.

### Simulation A

Simulation A was conducted to take into account the uncertainty around the reported parameter estimates for incubation period, generation time and serial interval. Where we could extract enough information from the publications, we sampled from distributions capturing the 95% confidence intervals around the mean and standard deviation for each reported distribution (n=1000). We converted these samples to the relevant parameters for the distribution (e.g. shape and rate for a gamma distribution) and simulated distributions using these parameters (n= 1000). The incubation period sample was subtracted from each generation time or serial interval sample to give a resultant distribution indicating transmission time relative to onset of symptoms. The resultant 1,000,000 samples were resampled with replacement (n=1000 samples from each of 10,000 repeats) and 95% confidence intervals from bootstrapping were calculated.

As there are known drivers of variation of generation time and serial interval [34], and therefore of transmission time relative to symptom onset estimated based upon them, estimates were plotted and summarised individually to show this variation. Estimates were also grouped, plotted and summarised at the level of source location for the contact tracing data that they were inferred from. Simple unweighted pooled estimates of transmission time relative to symptom onset based on serial interval and generation time were presented for interpretation in the context of the variation between the individual results.

### Simulation B

Simulation B incorporated data from every study that reported enough information to simulate a full distribution for serial interval or generation time but did not take uncertainty around the parameter estimates into account (as this was not reported in all of the studies). We simulated data (n = 100,000 samples) for incubation period and generation time or serial interval from each study. The distributions were plotted, summarised and cross-checked against the summary statistics and plots reported in the papers. The incubation period sample was subtracted from the generation time or serial interval sample to give a resultant distribution indicating transmission time relative to onset of symptoms. This was plotted and summarised and the proportion of samples of transmission times relative to symptom onset that were negative (indicating transmission before symptoms was reported) was calculated. This simulation was repeated 20,000 times to explore the uncertainty from within the simulation. As with Simulation A, summaries at estimate, source location and pooled level were reported. Unlike Simulation A, confidence intervals were based only on the variation from within the simulation.

## 4. Results

### Included studies

Of the 19 studies reporting serial interval or generation time included in our rapid systematic review [34], 17 were included in this study. We excluded the study [54] as it defined the start of the exposure window for the infectee as the time of symptom onset in the infector, excluding the possibility of transmission before symptom onset. We excluded one further study, [20], pending clarification from the authors, as we could not replicate the distribution described for serial interval. Table 2 lists the estimates considered for inclusion. Figure 2 describes the data available for 28 estimates for serial interval or generation time from the 17 publications that were included in this study.

**Table 2:**
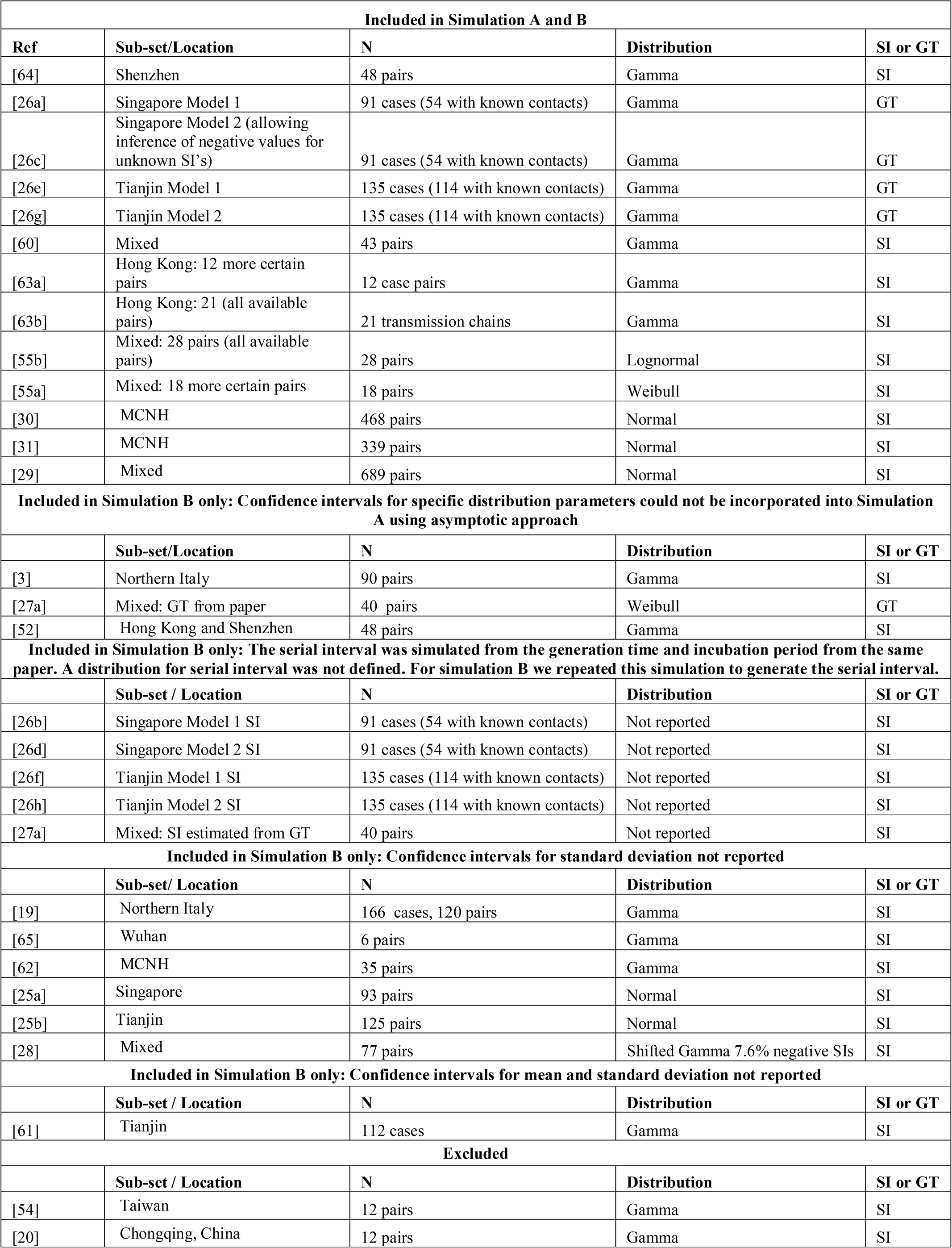
A summary of all generation time and serial interval estimates included in our rapid systematic review [34] considered for inclusion in this study, those included in both Simulations A and B, those included in Simulation A only, and those excluded. SI = Serial interval. GT = Generation time. Ref = reference.

**Figure 2:**
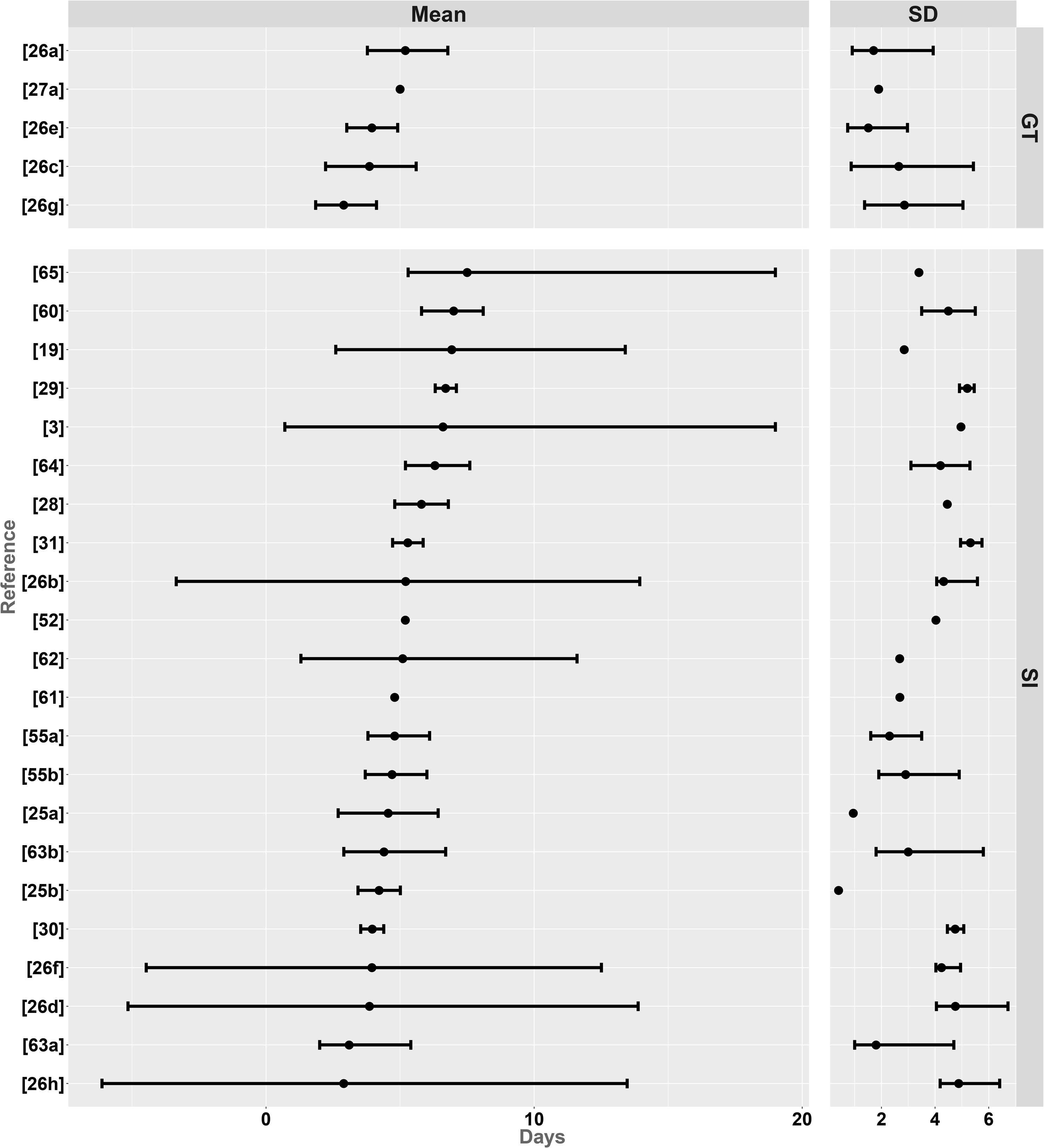
Generation time (GT, upper panels) and serial interval (SI, lower panels) estimates from the publications included in this study. Mean (left panels) and Standard Deviation (SD, right panels) are shown. Circles represent mean estimates and bars represent 95% confidence intervals.

### Simulation A results

Studies that reported 95% confidence intervals for both the mean and standard deviation of the generation time or serial interval could be incorporated into Simulation A (Table 2). These included four generation time estimates and nine serial interval estimates from eight different studies. Four of these estimates came from mixed locations, two each came from Hong Kong, Mainland China excluding Hubei, Singapore and Tianjin, and one came from Shenzhen. The uncertainty captured by the simulation was slightly less than that reported in the source publications for serial interval and generation time. (Supplementary Table 1).

Table 3 summarises transmission time relative to symptom onset for each of the 13 estimates included in Simulation A. These ranged from a mean of 2.91 (95% CI: 3.18, 2.64) days before symptom onset to 1.20 (95% CI: 0.86, 1.55) days after symptom onset. Simple unweighted pooling of estimates of transmission time relative to symptom onset based on serial interval estimates resulted in a mean of 0.60 days before symptom onset (95% CI: 3.01 days before, 1.81 days after). Pooling based on generation time estimates gave a mean of 1.83 (95% CI: 3.48, 0.17) days before symptom onset.

**Table 3:** Summary statistics for transmission time relative to symptom onset and proportion pre-symptomatic transmission estimated from Simulation A. The studies are ordered first by whether they are based on generation time (GT) or serial interval (SI), and then by median transmission time relative to symptoms. A negative number means days before symptom onset. MCNH = Mainland China excluding Hubei province. Mixed = pooled data from multiple countries. SD = Standard deviation. Ref = reference number corresponding to Table 2 and main text. Confidence intervals are based incorporating measures of uncertainty from the source publications into the simulation.

| Ref | Location | SI or GT | Percentage pre-symptomatic transmission (95% CI) | Mean (95% CI) | SD (95% CI) | 2.5th percentile (95% CI) | 25th percentile (95% CI) | Median (95% CI) | 75th percentile (95% CI) | 97.5th percentile (95% CI) |
| --- | --- | --- | --- | --- | --- | --- | --- | --- | --- | --- |
| [26a] | Singapore | GT | 52.1 (49.0, 55.2) | -0.57 (-0.80, -0.34) | 3.71 (3.44, 3.98) | -9.03 (-10.17, -7.89) | -2.48 (-2.80, -2.16) | -0.16 (-0.40, 0.08) | 1.79 (1.55, 2.02) | 5.55 (4.96, 6.15) |
| [26e] | Tianjin | GT | 69.3 (66.4, 72.1) | -1.84 (-2.06, -1.63) | 3.54 (3.27, 3.81) | -10.13 (-11.27, -8.99) | -3.60 (-3.91, -3.28) | -1.38 (-1.61, -1.15) | 0.43 (0.22, 0.64) | 3.76 (3.25, 4.27) |
| [26c] | Singapore | GT | 71.1 (68.3, 73.9) | -1.98 (-2.25, -1.71) | 4.35 (3.99, 4.71) | -10.87 (-12.00, -9.73) | -4.31 (-4.63, -3.98) | -1.91 (-2.17, -1.64) | 0.41 (0.11, 0.71) | 6.44 (5.28, 7.61) |
| [26g] | Tianjin | GT | 80.6 (78.1, 83.0) | -2.91 (-3.18, -2.64) | 4.39 (4.01, 4.77) | -11.70 (-12.82, -10.58) | -5.19 (-5.50, -4.87) | -2.91 (-3.15, -2.66) | -0.71 (-1.01, -0.40) | 6.03 (4.70, 7.35) |
| [29] | Mixed | SI | 42.8 (39.8, 45.9) | 0.90 (0.52, 1.28) | 6.11 (5.81, 6.40) | -11.59 (-12.81, -10.37) | -3.01 (-3.54, -2.48) | 1.08 (0.62, 1.53) | 5.03 (4.54, 5.52) | 12.31 (11.38, 13.23) |
| [60] | Mixed | SI | 43.4 (40.4, 46.5) | 1.20 (0.86, 1.55) | 5.54 (5.19, 5.88) | -8.86 (-9.96, -7.76) | -2.20 (-2.56, -1.83) | 0.76 (0.40, 1.12) | 4.24 (3.76, 4.72) | 13.38 (11.92, 14.84) |
| [64] | Shenzhen | SI | 48.6 (45.5, 51.7) | 0.51 (0.18, 0.84) | 5.28 (4.95, 5.62) | -9.31 (-10.46, -8.17) | -2.67 (-3.02, -2.32) | 0.15 (-0.19, 0.50) | 3.41 (2.95, 3.86) | 11.97 (10.59, 13.35) |
| [31] | MCNH | SI | 51.9 (48.8, 55.0) | -0.47 (-0.86, -0.08) | 6.28 (5.97, 6.58) | -13.24 (-14.50, -11.98) | -4.51 (-5.04, -3.97) | -0.30 (-0.77, 0.18) | 3.77 (3.26, 4.28) | 11.29 (10.34, 12.24) |
| [55a] | Mixed | SI | 56.2 (53.1, 59.3) | -0.78 (-1.06, -0.50) | 4.46 (3.45, 5.46) | -10.10 (-11.22, -8.97) | -3.44 (-3.80, -3.08) | -0.66 (-0.99, -0.33) | 2.18 (1.82, 2.53) | 7.26 (6.63, 7.90) |
| [55b] | Mixed | SI | 58.8 (55.7, 61.9) | -0.81 (-1.11, -0.50) | 4.81 (4.29, 5.33) | -10.23 (-11.37, -9.08) | -3.59 (-3.94, -3.24) | -0.92 (-1.24, -0.61) | 1.96 (1.58, 2.35) | 8.68 (7.58, 9.78) |
| [30] | MCNH | SI | 61.7 (58.6, 64.7) | -1.84 (-2.19, -1.48) | 5.75 (5.47, 6.04) | -13.69 (-14.89, -12.48) | -5.47 (-5.96, -4.99) | -1.64 (-2.07, -1.21) | 2.05 (1.59, 2.51) | 8.81 (7.97, 9.64) |
| [63b] | Hong Kong | SI | 65.1 (62.1, 68.1) | -1.42 (-1.70, -1.13) | 4.54 (4.20, 4.89) | -10.51 (-11.64, -9.37) | -3.93 (-4.26, -3.60) | -1.43 (-1.72, -1.15) | 1.10 (0.77, 1.43) | 7.67 (6.46, 8.88) |
| [63a] | Hong Kong | SI | 79.3 (76.8, 81.8) | -2.69 (-2.93, -2.45) | 3.83 (3.46, 4.20) | -11.16 (-12.32, -9.99) | -4.62 (-4.94, -4.31) | -2.36 (-2.60, -2.13) | -0.40 (-0.64, -0.16) | 3.85 (3.08, 4.62) |
| Unweighted pooling |  | GT | 68.3 (47.9, 88.6) | -1.83 (-3.48, -0.17) | 4.00 (3.19, 4.80) | -10.43 (-12.66, -8.20) | -3.89 (-5.86, -1.92) | -1.59 (-3.54, 0.37) | 0.48 (-1.27, 2.23) | 5.45 (3.22, 7.67) |
|  |  | SI | 56.4 (34.9, 78.0) | -0.60 (-3.01, 1.81) | 5.18 (3.58, 6.77) | -10.96 (-14.22, -7.70) | -3.72 (-5.67, -1.76) | -0.59 (-2.72, 1.54) | 2.59 (-0.55, 5.74) | 9.47 (3.76, 15.18) |

Proportion of pre-symptomatic transmission calculated from the 13 individual estimates ranged from 42.8% (95% CI:39.8%, 45.9%) to 80.6% (78.1%, 83.0%) (Table 3).

From unweighted pooling of serial interval and generation time estimates respectively, the proportion of pre-symptomatic transmission was estimated to be 56.4% (95% CI: 34.9%, 78.0%) and 68.3% (95% CI: 47.9%, 88.6%) (Table 3).

Simple unweighted pooling at source location level resulted in mean transmission time relative to symptoms ranging from 2.38 (95% CI: 3.45, 1.30) days before symptom onset in Tianjin to a mean of 0.51 (95% CI: 0.18, 0.84) days after symptom onset in Shenzhen. Proportion of pre-symptomatic transmission ranged from 48.6% (95% CI: 45.5%, 51.7%) in Shenzhen to 74.9% (95% CI: 63.5%, 86.3%) in Tianjin (Table 4).

**Table 4:** Summary statistics for transmission time relative to symptom onset and proportion pre-symptomatic transmission estimated from Simulation A. MCNH = Mainland China excluding Hubei province. N Italy = Northern Italy. Mixed = data came from multiple countries. SD = Standard deviation. Confidence intervals in Simulation A are based on incorporating measures of uncertainty from the source publications into the simulation.

| Location | Percentage pre-symptomatic transmission | Mean (95% CI) | SD (95% CI) | 2.5th percentile (95% CI) | 25th percentile (95% CI) | Median (95% CI) | 75th percentile (95% CI) | 97.5th percentile (95% CI) |
| --- | --- | --- | --- | --- | --- | --- | --- | --- |
| Shenzhen | 48.6 (45.5, 51.7) | 0.51 (0.18, 0.84) | 5.28 (4.95, 5.62) | -9.31 (-10.46, -8.17) | -2.67 (-3.02, -2.32) | 0.15 (-0.19, 0.50) | 3.41 (2.95, 3.86) | 11.97 (10.59, 13.35) |
| Mixed | 50.3 (35.8, 64.9) | 0.13 (-1.72, 1.98) | 5.23 (3.84, 6.62) | -10.19 (-12.41, -7.98) | -3.06 (-4.19, -1.92) | 0.06 (-1.68, 1.80) | 3.35 (0.74, 5.97) | 10.41 (5.36, 15.46) |
| MCNH | 56.8 (46.8, 66.8) | -1.15 (-2.55, 0.24) | 6.01 (5.42, 6.61) | -13.46 (-14.77, -12.15) | -4.99 (-6.07, -3.91) | -0.97 (-2.36, 0.43) | 2.91 (1.16, 4.67) | 10.05 (7.45, 12.64) |
| Singapore | 61.6 (42.7, 80.5) | -1.2 (-2.68, 0.13) | 4.03 (3.33, 4.73) | -9.95 (-12.08, -7.82) | -3.39 (-5.21, -1.57) | -1.04 (-2.77, 0.70) | 1.10 (-0.27, 2.47) | 6.00 (4.72, 7.27) |
| Hong Kong | 72.2 (58.0, 86.3) | -2.05 (-3.33, -0.78) | 4.19 (3.40, 4.97) | -10.83 (-12.15, -9.52) | -4.28 (-5.03, -3.52) | -1.90 (-2.84, -0.95) | 0.35 (-1.15, 1.85) | 5.76 (1.88, 9.64) |
| Tianjin | 74.9 (63.5, 86.3) | -2.38 (-3.45, -1.30) | 3.97 (3.07, 4.86) | -10.91 (-12.83, -9.00) | -4.39 (-5.98, -2.80) | -2.14 (-3.66, -0.63) | -0.14 (-1.29, 1.01) | 4.89 (2.46, 7.33) |

Further details of standard deviation, 2.5^th^, 25^th^, 50^th^, 75^th^ and 97.5^th^ quantiles of the distributions described above, and their 95% confidence intervals are shown in Tables 3 and 4.

### Simulation B results

A total of 28 estimates from 17 studies were included for Simulation B. Transmission time relative to symptom onset estimates were based on five estimates of generation times and 23 estimates of serial interval. Several studies generated more than one estimate. This was due to separate estimates for different locations [25,26], different models used to infer generation time [26], sub-setting of data depending on confidence in transmission pair identification and exposure windows [52,55], and estimation of both generation times and serial intervals from the same papers [26,27]. Of the two models used in [26], one only allowed positive serial intervals to be inferred for missing data whereas a second model allowed negative serial intervals for missing data.

Table 5 shows the counts of estimates and studies that came from specific locations or mixed sources under nine different data source categories (Mixed sources, Tianjin, Singapore, Mainland China excluding Hubei, Hong Kong, northern Italy, pooled data from Hong Kong and Shenzhen, and Wuhan).

**Table 5:** A summary of the counts (N) of studies and estimates used in Simulation B coming from specific locations, or mixed sources.

| Location / data source | Count of estimates | Count of studies | References |
| --- | --- | --- | --- |
| Mixed | 7 | 5 | [27–29,55,61] |
| Tianjin | 6 | 3 | [25,26,62] |
| Singapore | 5 | 2 | [25,26] |
| Mainland China excluding Hubei | 3 | 3 | [30,31,63] |
| Hong Kong | 2 | 1 | [64] |
| Northern Italy | 2 | 2 | [3,19] |
| Hong Kong and Shenzhen | 1 | 1 | [52] |
| Shenzhen | 1 | 1 | [65] |
| Wuhan | 1 | 1 | [66] |
| **Total** | **28** | **19** |  |

Results for Simulation B, incorporating a relatively broader range of studies, were similar to those for Simulation A. Figure 3 and Table 6 show the variation in transmission time relative to symptom onset amongst the 28 estimates ranging from a mean (median) of 2.92 (2.91) days before symptom onset to 1.72 (1.69) days after symptom onset. Proportion of pre-symptomatic transmission associated with the 28 different estimates ranged from 33.5% to 80.7%.

**Figure 3:**
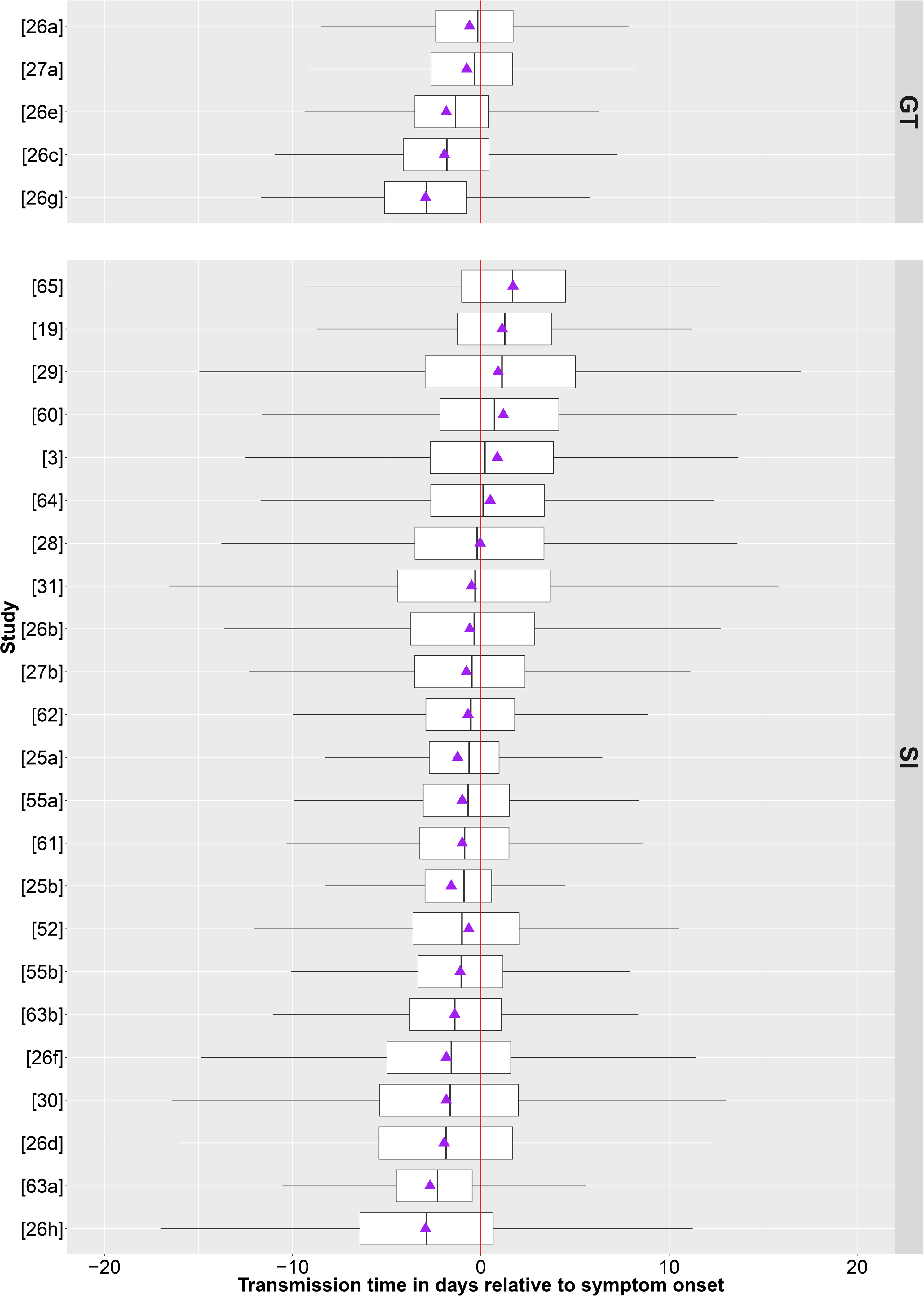
A boxplot showing time of transmission relative to onset of symptoms inferred from simulations of incubation period and generation time (GT) (n = 5 estimates) or serial interval (SI) (n = 23 estimates). The purple triangles represent the mean of the simulated samples. The vertical red line represents onset of symptoms. The numbers on the left axis are the reference numbers for each estimate (described in Table 2).

**Table 6:** Summary statistics based on Simulation B for transmission time relative to symptom onset and proportion pre-symptomatic transmission (PPS) for each of 5 estimates based on generation time (GT) and 23 estimates based on serial interval (SI). The studies are ordered first by whether they are based on generation time or serial interval, and then by median transmission time relative to symptoms, to correspond to Figure 3. A negative number means days before symptom onset. MCNH = Mainland China excluding Hubei province. Mixed = SI or GT was calculated from pooled data from multiple countries. HKSZ = pooled data from Hong Kong and Shenzhen. SD = Standard deviation. Figure 2 ref = Reference number corresponding to Table 2, Figure 3 and main text. Confidence intervals are based on the variation between 20,000 repeat simulations.

| Reference | Location (description) | N for study | Based on | % pre-symptomatic transmission | Mean | Standard deviation | 2.5th percentile | 25th percentile | Median | 75th percentile | 97.5th percentile |
| --- | --- | --- | --- | --- | --- | --- | --- | --- | --- | --- | --- |
| [26a] | Singapore | 91 (54) cases | GT | 52.14 (51.83, 52.46) | -0.59 (-0.61, -0.57) | 3.53 (3.50, 3.55) | -8.87 (-8.99, -8.76) | -2.38 (-2.41, -2.35) | -0.16 (-0.18, -0.13) | 1.71 (1.69, 1.73) | 5.13 (5.09, 5.18) |
| [27a] | Mixed | 40 pairs | GT | 54.14 (53.83, 54.45) | -0.74 (-0.76, -0.71) | 3.64 (3.61, 3.66) | -9.19 (-9.30, -9.07) | -2.66 (-2.69, -2.63) | -0.33 (-0.35, -0.30) | 1.70 (1.68, 1.73) | 5.16 (5.12, 5.20) |
| [26e] | Tianjin | 135 (114) cases | GT | 69.42 (69.14, 69.71) | -1.83 (-1.85, -1.81) | 3.43 (3.41, 3.46) | -10.01 (-10.12, -9.89) | -3.53 (-3.56, -3.50) | -1.35 (-1.38, -1.33) | 0.42 (0.39, 0.44) | 3.58 (3.53, 3.62) |
| [26c] | Singapore Model 2 | 93 (54) cases | GT | 70.89 (70.61, 71.18) | -1.93 (-1.95, -1.90) | 4.06 (4.04, 4.09) | -10.63 (-10.74, -10.52) | -4.13 (-4.16, -4.09) | -1.81 (-1.83, -1.78) | 0.43 (0.40, 0.46) | 5.99 (5.91, 6.08) |
| [26g] | Tianjin Model 2 | 137 (114) cases | GT | 80.70 (80.45, 80.94) | -2.92 (-2.95, -2.90) | 4.18 (4.15, 4.21) | -11.61 (-11.72, -11.50) | -5.13 (-5.16, -5.10) | -2.88 (-2.90, -2.85) | -0.73 (-0.76, -0.70) | 5.74 (5.63, 5.84) |
| [65] | Wuhan | 6 pairs | SI | 33.47 (33.18, 33.76) | 1.72 (1.69, 1.75) | 4.59 (4.56, 4.62) | -7.63 (-7.74, -7.52) | -1.00 (-1.04, -0.97) | 1.69 (1.66, 1.73) | 4.50 (4.46, 4.54) | 10.89 (10.80, 10.98) |
| [19] | N Italy | 120 pairs | SI | 36.20 (35.90, 36.49) | 1.15 (1.13, 1.18) | 4.20 (4.17, 4.22) | -7.81 (-7.92, -7.70) | -1.23 (-1.26, -1.20) | 1.29 (1.26, 1.32) | 3.76 (3.73, 3.79) | 9.11 (9.03, 9.19) |
| [29] | Mixed | 689 pairs | SI | 42.69 (42.39, 43.00) | 0.92 (0.88, 0.95) | 6.04 (6.02, 6.07) | -11.48 (-11.60, -11.35) | -2.96 (-3.02, -2.91) | 1.09 (1.05, 1.14) | 5.01 (4.96, 5.06) | 12.27 (12.18, 12.36) |
| [60] | Mixed | 43 pairs | SI | 43.22 (42.91, 43.53) | 1.22 (1.18, 1.25) | 5.45 (5.42, 5.49) | -8.80 (-8.91, -8.69) | -2.14 (-2.18, -2.11) | 0.77 (0.73, 0.81) | 4.23 (4.18, 4.28) | 13.29 (13.15, 13.43) |
| [3] | N Italy | 90 pairs | SI | 47.77 (47.46, 48.09) | 0.90 (0.86, 0.93) | 5.78 (5.74, 5.81) | -9.30 (-9.41, -9.19) | -2.66 (-2.70, -2.63) | 0.26 (0.22, 0.30) | 3.93 (3.88, 3.98) | 14.12 (13.96, 14.28) |
| [64] | Shenzhen | 48 pairs | SI | 48.68 (48.37, 49.00) | 0.52 (0.48, 0.55) | 5.21 (5.18, 5.24) | -9.24 (-9.35, -9.13) | -2.63 (-2.66, -2.59) | 0.14 (0.11, 0.18) | 3.39 (3.34, 3.43) | 11.95 (11.82, 12.08) |
| [28] | MCNH | 339 pairs | SI | 51.54 (51.23, 51.85) | -0.02 (-0.06, 0.01) | 5.44 (5.41, 5.47) | -10.49 (-10.60, 10.38) | -3.49 (-3.53, -3.45) | -0.19 (-0.23, -0.16) | 3.34 (3.29, 3.38) | 11.18 (11.07, 11.29) |
| [31] | Mixed | 77 pairs | SI | 52.15<br>(51.84, 52.46) | -0.49<br>(-0.53, -0.46) | 6.15<br>(6.12, 6.18) | -13.07<br>(-13.20, -12.95) | -4.45<br>(-4.50, -4.40) | -0.32<br>(-0.37, -0.28) | 3.67<br>(3.62, 3.72) | 11.08<br>(10.98, 11.17) |
| [26b] | Singapore | 92<br>(54) cases | SI | 53.02<br>(52.71, 53.33) | -0.59<br>(-0.62, -0.56) | 5.30<br>(5.28, 5.33) | -11.78<br>(-11.90, -11.66) | -3.80<br>(-3.85, -3.76) | -0.37<br>(-0.41, -0.33) | 2.87<br>(2.83, 2.91) | 9.35<br>(9.26, 9.44) |
| [27b] | Mixed | 40 pairs | SI | 54.30<br>(53.99, 54.61) | -0.74<br>(-0.77, -0.71) | 4.67<br>(4.65, 4.70) | -10.82<br>(-10.94, -10.71) | -3.50<br>(-3.54, -3.46) | -0.46<br>(-0.49, -0.43) | 2.36<br>(2.32, 2.39) | 7.70<br>(7.63, 7.77) |
| [62] | MCNH | 35 pairs | SI | 56.19<br>(55.88, 56.50) | -0.67<br>(-0.70, -0.65) | 4.09<br>(4.06, 4.11) | -9.46<br>(-9.57, -9.35) | -2.93<br>(-2.96, -2.90) | -0.53<br>(-0.56, -0.50) | 1.81<br>(1.78, 1.84) | 7.11<br>(7.04, 7.19) |
| [25a] | Singapore | 93 pairs | SI | 59.57<br>(59.26, 59.87) | -1.22<br>(-1.24, -1.20) | 3.23<br>(3.20, 3.25) | -9.20<br>(-9.31, -9.09) | -2.74<br>(-2.77, -2.71) | -0.62<br>(-0.64, -0.60) | 0.98<br>(0.96, 1.00) | 3.25<br>(3.22, 3.27) |
| [55a] | Mixed (n=18) | 18 pairs | SI | 58.22<br>(57.91, 58.53) | -0.98<br>(-1.01, -0.96) | 3.85<br>(3.82, 3.87) | -9.62<br>(-9.74, -9.51) | -3.09<br>(-3.12, -3.05) | -0.69<br>(-0.72, -0.66) | 1.53<br>(1.50, 1.56) | 5.70<br>(5.64, 5.75) |
| [61] | Tianjin | 112 cases | SI | 59.96<br>(59.66, 60.27) | -0.98<br>(-1.01, -0.96) | 4.10<br>(4.07, 4.12) | -9.77<br>(-9.88, -9.66) | -3.24<br>(-3.27, -3.21) | -0.85<br>(-0.88, -0.83) | 1.48<br>(1.45, 1.51) | 6.90<br>(6.81, 6.98) |
| [25b] | Tianjin | 125 pairs | SI | 64.88<br>(64.59, 65.18) | -1.56<br>(-1.58, -1.54) | 3.11<br>(3.08, 3.13) | -9.41<br>(-9.52, -9.29) | -2.96<br>(-2.99, -2.93) | -0.90<br>(-0.92, -0.88) | 0.59<br>(0.57, 0.60) | 2.42<br>(2.41, 2.44) |
| [52] | HKSZ | 48 pairs | SI | 59.34<br>(59.04, 59.65) | -0.61<br>(-0.64, -0.58) | 5.05<br>(5.02, 5.09) | -10.12<br>(-10.23, -10.01) | -3.56<br>(-3.59, -3.53) | -0.98<br>(-1.01, -0.94) | 2.07<br>(2.02, 2.11) | 10.60<br>(10.46, 10.74) |
| [55b] | Mixed (n=28) | 28 pairs | SI | 62.63<br>(62.33, 62.93) | -1.08<br>(-1.11, -1.06) | 4.23<br>(4.20, 4.26) | -9.84<br>(-9.95, -9.73) | -3.34<br>(-3.37, -3.31) | -1.03<br>(-1.05, -1.00) | 1.19<br>(1.16, 1.22) | 7.50<br>(7.39, 7.61) |
| [63b] | Hong Kong (n=21) | 21 pairs | SI | 65.24<br>(64.94, 65.53) | -1.38<br>(-1.41, -1.36) | 4.30<br>(4.27, 4.33) | -10.30<br>(-10.41, -10.19) | -3.78<br>(-3.81, -3.74) | -1.38<br>(-1.40, -1.35) | 1.08<br>(1.04, 1.11) | 7.32<br>(7.22, 7.41) |
| [30] | MCNH | 468 pairs | SI | 63.17<br>(62.87, 63.47) | -1.83<br>(-1.86, -1.80) | 5.24<br>(5.21, 5.27) | -12.93<br>(-13.05, -12.81) | -4.99<br>(-5.03, -4.94) | -1.60<br>(-1.63, -1.56) | 1.59<br>(1.55, 1.63) | 7.96<br>(7.87, 8.05) |
| [26f] | Tianjin | 136<br>(114) cases | SI | 61.74<br>(61.45, 62.04) | -1.82<br>(-1.86, -1.79) | 5.66<br>(5.63, 5.69) | -13.53<br>(-13.65, -13.41) | -5.42<br>(-5.47, -5.37) | -1.63<br>(-1.67, -1.59) | 2.01<br>(1.97, 2.06) | 8.72<br>(8.63, 8.80) |
| [26d] | Singapore Model 2 | 94<br>(54) cases | SI | 63.91<br>(63.61, 64.20) | -1.93<br>(-1.96, -1.89) | 5.67<br>(5.64, 5.70) | -13.51<br>(-13.63, -13.39) | -5.43<br>(-5.48, -5.39) | -1.85<br>(-1.89, -1.81) | 1.68<br>(1.64, 1.72) | 9.18<br>(9.07, 9.28) |
| [63a] | Hong Kong (n=12) | 12 pairs | SI | 79.90<br>(79.65, 80.15) | -2.68<br>(-2.71, -2.66) | 3.57<br>(3.54, 3.59) | -10.97<br>(-11.08, -10.85) | -4.49<br>(-4.52, -4.46) | -2.29<br>(-2.31, -2.27) | -0.44<br>(-0.47, -0.42) | 3.37<br>(3.31, 3.42) |
| [26h] | Tianjin Model 2 | 138<br>(114) cases | SI | 71.10<br>(70.82, 71.38) | -2.92<br>(-2.96, -2.89) | 5.76<br>(5.72, 5.79) | -14.51<br>(-14.63, -14.39) | -6.47<br>(-6.51, -6.42) | -2.91<br>(-2.95, -2.87) | 0.64<br>(0.59, 0.68) | 8.61<br>(8.49, 8.73) |
| Simple pooled estimate of GT based transmission time relative to symptom onset |  |  |  | 65.46<br>(65.27, 65.65) | -1.60<br>(-1.62, -1.58) | 3.88<br>(3.85, 3.90) | -10.25<br>(-10.34, -10.15) | -3.71<br>(-3.73, -3.68) | -1.31<br>(-1.33, -1.29) | 0.88<br>(0.87, 0.90) | 5.12<br>(5.09, 5.14) |
| Simple pooled estimate of SI based transmission time relative to symptom onset |  |  |  | 56.04<br>(55.89, 56.19) | -0.66<br>(-0.68, -0.64) | 5.05<br>(5.03, 5.06) | -11.10<br>(-11.17, -11.03) | -3.51<br>(-3.54, -3.49) | -0.62<br>(-0.64, -0.60) | 2.21<br>(2.20, 2.23) | 9.60<br>(9.58, 9.63) |

Table 6 also shows the summary statistics for a simple unweighted pooling of the estimates based on serial interval and the five estimates based on generation time. The mean (median) time of transmission for the pooled samples based on serial interval was 0.66 (0.62) days before symptoms and pre-symptomatic transmission was estimated to be 56.0%. The pooling of five generation time based estimates for transmission time relative to symptom onset gave a mean (median) estimate of 1.60 (1.31) days before symptom onset and 65.5% pre-symptomatic transmission.

Figure 4 and Table 7 show the variation in transmission relative to symptom onset amongst estimates based on the nine different source categories ranging from a mean (median) of 2.05 (1.90) days before symptom onset for Hong Kong to 1.72 (1.68) days after symptom onset for Wuhan. Proportion of pre-symptomatic transmission amongst the different data source categories ranged from 33.5% in Wuhan to 72.7% in Hong Kong.

**Figure 4:**
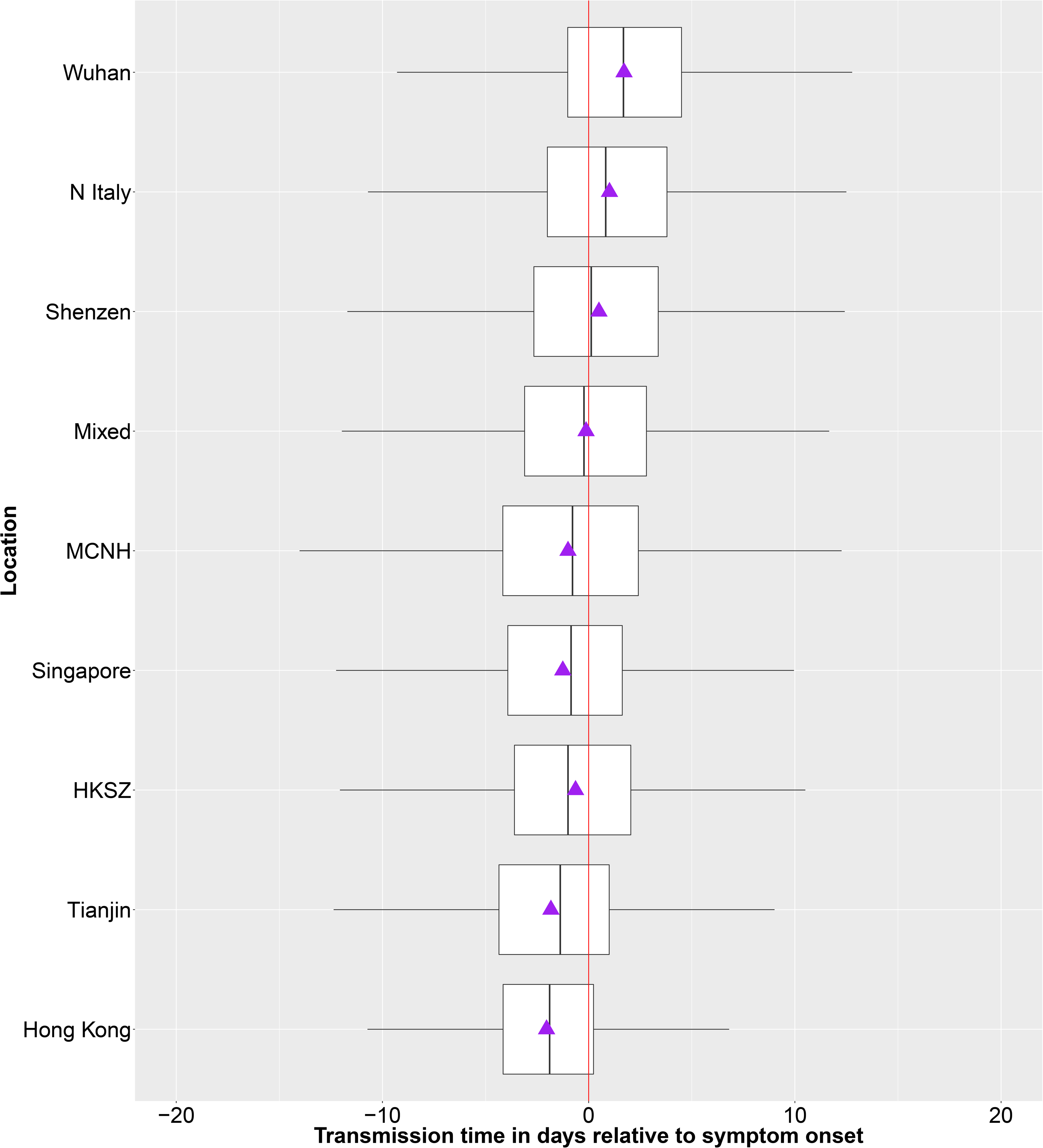
A boxplot showing time of transmission relative to onset of symptoms inferred from simulations from incubation period and serial interval (n = 23 estimates). The purple triangles represent the mean of the simulated samples. The vertical red line represents onset of symptoms. The simulated samples were pooled according to the source location of the original data upon which estimates were based. MCNH = Mainland China excluding Hubei province. N Italy = Northern Italy. HKSZ = pooled data from Hong Kong and Shenzhen. Mixed = data came from multiple countries.

**Table 7:** Summary statistics from Simulation B for transmission time relative to symptom onset and proportion pre-symptomatic transmission (PPS) for simulated samples, based on 23 estimates of serial interval, pooled according to the source location of the original data upon which estimates were based. MCNH = Mainland China excluding Hubei province. N Italy = Northern Italy. Mixed = data came from multiple countries. The studies are ordered by median transmission time relative to symptoms, to correspond to Figure 4. SD = Standard deviation.

| Location | Count of estimates | Proportion pre-symptomatic transmission | Mean | SD | 2.5th percentile | 25th percentile | Median | 75th percentile | 97.5th percentile |
| --- | --- | --- | --- | --- | --- | --- | --- | --- | --- |
| Wuhan | 1 | 33.5% | 1.72 | 4.62 | -7.63 | -1.02 | 1.68 | 4.53 | 10.96 |
| N Italy | 2 | 42.1% | 1.01 | 5.05 | -8.66 | -2.00 | 0.83 | 3.81 | 11.76 |
| Shenzhen | 1 | 48.9% | 0.50 | 5.22 | -9.26 | -2.65 | 0.12 | 3.40 | 11.90 |
| Mixed | 6 | 52.2% | -0.13 | 5.09 | -10.29 | -3.13 | -0.23 | 2.81 | 10.49 |
| MCNH | 3 | 56.8% | -1.01 | 5.41 | -12.48 | -4.19 | -0.80 | 2.40 | 9.35 |
| Singapore | 3 | 58.8% | -1.25 | 4.89 | -11.95 | -3.97 | -0.87 | 1.64 | 8.27 |
| HKSZ | 1 | 59.3% | -0.62 | 5.06 | -10.16 | -3.59 | -0.96 | 2.07 | 10.64 |
| Tianjin | 4 | 64.9% | -1.84 | 4.72 | -12.31 | -4.40 | -1.40 | 0.99 | 7.09 |
| Hong Kong | 2 | 72.7% | -2.05 | 4.00 | -10.67 | -4.19 | -1.90 | 0.24 | 5.81 |

For simulation B, the uncertainty captured by 20,000 repeat simulations was small (Table 6), as would be expected for large samples in simulations from defined distributions.

## 5. Discussion

Our results show that transmission time ranged from mean of 2.92 days before symptom onset to 1.72 days after symptom onset. Simple unweighted pooling of the 23 estimates based on serial intervals resulted in a mean time of transmission of 0.66 days before symptoms. From this, it can be inferred that transmission of SARS-CoV-2 is most likely in the day before symptom onset. The estimates suggesting most pre-symptomatic transmission highlighted a mean transmission times almost 3 days before symptom onset. This is consistent with other estimates in the literature, ranging from 2.65 days before symptoms to 2.3 days after symptoms [25-29,32]. A study focussed on inferring infectiousness profile from 77 transmission pairs, reported that infectiousness started from 2.3 days before symptom onset and peaked at 0.7 days before symptom onset [28]. Our study, analysing all estimates from a variety of locations has produced a similar estimate (mean from pooled estimate of 0.67 days before symptom onset).

We estimated that the proportion of pre-symptomatic transmission ranged from 33.5% to 80.7% depending on the study analysed. Pooled serial interval based estimates suggested 56.0% pre-symptomatic transmission. Our source level analyses describe a trend in pre-symptomatic transmission ranging from the greatest proportion in Hong Kong (72.7%) to the smallest proportion in Wuhan (33.5%). The range in the proportion of pre-symptomatic transmission that we report is consistent with other studies [25,26,28,29,32] where reports of pre-symptomatic transmission range from 37% to >80%.

The wide variation in estimates of the proportion of pre-symptomatic transmission and transmission time relative to symptom onset is expected, as the observed transmission events depend on contact rates between infectious and susceptible people as well as the natural course of infectiousness. The variation in observed transmission events is manifested by variations in serial interval and generation time, and therefore in transmission relative to symptom onset. Griffin et al. [34] discuss drivers of this variation and the limitations in different approaches to estimating generation time and serial interval from transmission pairs. If people are quarantined once their symptoms become apparent, a greater proportion transmission will be pre-symptomatic. Whilst data relating to the early stages of the COVID-19 outbreak in Wuhan amount to only six pairs, we see a trend towards lower proportions of pre-symptomatic transmission in this context, as well as in the early stages of the outbreaks in Italy, possibly corresponding to relatively more transmission from symptomatic people in the early stages of disease incursion. Three other studies also highlight this contrast in the proportion of pre-symptomatic transmission in a different context. Zhao et al. [52] show that serial interval became shorter in Hong Kong and Shenzhen as time elapsed from initial cases, and suggest that this is due to increasing effectiveness of quarantining people with symptoms over time. Zhang [32] contrasts a mean transmission time of 2.3 days after symptoms in the early stages of the Wuhan outbreak (with relatively fewer quarantine measures) to a mean transmission time of 2.4 days before symptom onset amongst imported cases outside Wuhan. A study in Shenzhen [64] highlighted the valuable information that can be inferred from knowledge of when transmission occurred and when the infector was isolated. Serial interval was much shorter when the infectors were isolated within two days of symptom onset compared to those isolated 3-5 days after symptom onset. However, lack of isolation beyond five days made little difference, suggesting that there may be relatively more potential for transmission before compared to after five days of symptom onset. A further analysis [56] highlights the difference in pre-symptomatic transmission in Shenzhen between a subset with accelerated case isolation (46% pre-symptomatic transmission) one without (23%).

Despite this understandable variation in estimates from different contexts, our work consistently suggests the potential for pre-symptomatic transmission. A person presenting with COVID-19 symptoms has potentially been infectious to others for several days. Therefore, in absence of severe social distancing measures, extremely effective and rapid contact tracing and quarantine will be required to control the spread of COVID-19.

This potential of the COVID-19 spread to be too fast to be controlled by conventional contact tracing been highlighted with Ferretti et al.’s model [27]. This model suggests that pre-symptomatic transmission alone can account for a basic reproductive number of 0.9 (47% of the overall reproductive basic number), almost enough to sustain an epidemic on its own. However, this estimate may be influenced by the low level of asymptomatic infectiousness (10% relative to a symptomatic case) assumed by that model. This uncertainty highlights the need for transmission from asymptomatically infected people to be more fully understood, and to be considered as having potentially distinct characteristics compared to the pre-symptomatic transmission that we report on in this paper.

We used a straightforward approach of simulating large numbers of samples from both the incubation period distribution from our meta-analysis [33] and the distributions of 28 serial interval or generation time estimates from our rapid systematic review [34], and subtracting the incubation period samples from the serial interval or generation time samples to give a resultant distribution of transmission time relative to symptom onset. This methodology is similar to that applied by Tindale et al. and Ganyani et al. [25,26] to data from Singapore and Tianjin. An alternative method, using maximum likelihood estimation, was used by He et al. [28] and Zhang [32], and a Bayesian approach to estimate the probability of pre-symptomatic transmission was used by Ferretti et al. [27]. Ma et al. [29] estimated transmission time directly from manual interrogation of the data. Despite the variety of methodologies used to estimate transmission time, results of our analysis gave similar ranges to the six other studies that investigated pre-symptomatic transmission in different contexts [25-29,32].

Case reports [11-20] and virological studies [21-23] support the occurrence of pre-symptomatic transmission suggested by the quantitative approaches reported here and elsewhere [25-29,32]. Whilst samples testing positive by polymerase chain reaction (PCR) do not always fully correlate with infectiousness [24,57,58], relatively lower cycle threshold (CT) values suggest higher virus loads. Two studies [21,23] that reported pre-symptomatic PCR CT values included some relatively low values. A report of pre-symptomatic PCR positive samples in ten nursing home residents reported a mean time of 3 days from sampling to onset of symptoms. In addition, the isolation of live virus from upper respiratory samples very soon after patient presentation with symptoms has been reported [24].

### Limitations and strengths

Table 4 shows the potential overlap in contact tracing data that the studies we analysed are based on. Therefore, a limitation of our study is that all our data sources cannot be considered completely independent. We partially address this by grouping estimates by the source location of the contact-tracing data upon which they were based. Another challenge was to fully capture the uncertainty in estimates with a simulation study. Where enough information was available from the source publications, we incorporated measures of uncertainty into Simulation A, and reported a comparison of the uncertainty suggested in our simulation to that in the original estimates. A strength of our approach is that it builds a picture of pre-symptomatic transmission from a range of estimates in the literature, facilitates discussion for the drivers of variation between them, and highlights the consistent message that consideration of pre-symptomatic transmission is critical for COVID-19 control policy. The important insights into COVID-19 transmission gleaned from the studies that contributed to our analyses, that used publicly available transmission pair data, highlights the immense value of allowing public access to anonymised transmission pair data.

### Conclusion

Although contact rates between symptomatic infectious and susceptible people are likely to influence the proportion of pre-symptomatic transmission, our study highlights substantial potential for pre-symptomatic transmission of COVID-19 in a range of different contexts. Our work suggests that transmission of SARS-CoV-2 is most likely in the day before symptom onset, whereas estimates suggesting most pre-symptomatic transmission highlighted a mean transmission times almost 3 days before symptom onset. These findings highlight the urgent need for extremely rapid and effective case detection, contact tracing and quarantine measures if strict social distancing measures are to be eased.

## Data Availability

All data referred to in this manuscript are publicly available.

## Patient and public involvement statement

It was not appropriate or possible to involve patients or the public in the design, or conduct, or reporting, or dissemination plans of our research

## Author contributions

MC conceptualized the study, extracted parameter definitions from the literature, performed the analyses and drafted the manuscript. JG led the rapid systematic review upon which the generation time and serial interval simulations are based. CM led the meta-analysis upon which the incubation period simulations are based upon. AC, KH, KW and KOB performed systematic literature searches upon which the incubation period, generation time, serial interval and pre-symptomatic transmission information reported here are based upon. SM conceptualized, initiated and managed the overall project. All authors supplemented the literature review and assessed the literature. All authors contributed to the manuscript and reviewed it.

## Data sharing statement

The data and code used for the analyses described in this paper are available in the Github repository: https://github.com/miriamcasey/covid-19 presymptomatic project

## Acknowledgements

Dr. Jamie Tratalos and Mr. Guy McGrath of the Centre for Veterinary Epidemiology and Risk Analysis gave valuable feedback on this manuscript.

